# Association of Right Heart Structure and Function with Cognition and Brain MRI Measures in the Multi-Ethnic Study of Atherosclerosis

**DOI:** 10.1101/2024.08.15.24312008

**Authors:** Hooman Kamel, Susan R. Heckbert, Timothy M. Hughes, Christopher L. Schaich, Samuel N. Lockhart, Marcia Pescador Jimenez, W.T. Longstreth, Thomas R. Austin, R. Nick Bryan, Guray Erus, Dhivya Srinivasan, Ilya M. Nasrallah, Costantino Iadecola, Sanjiv J. Shah

## Abstract

**Background:** Recent studies in selected populations suggest that impaired venous return may affect brain health. We examined associations of right heart structure and function with cognition and MRI markers of brain health in a community-based cohort.

**Methods:** The Multi-Ethnic Study of Atherosclerosis (MESA) is a longitudinal cohort study of individuals 45 to 84 years of age who were free of clinically apparent cerebrovascular or cardiovascular disease at baseline in 2000-2002. From echocardiograms performed at Exam 6 during 2016-2018, we measured: right atrial pressure and end-systolic area; pulmonary artery systolic pressure; and right ventricular free wall strain, fractional area change, and end-diastolic area. Outcome variables were the Cognitive Assessment Screening Instrument, Digit Span, and Digit Symbol Coding tests and MRI-determined volumes of total white matter, total gray matter, and white matter hyperintensity -- as well as fractional anisotropy. Cognitive measures were obtained at Exam 6, and MRI measures, within a median of 18 months of echocardiograms. Covariates were demographics, *APOE*-ε4 allele status, vascular risk factors, and echocardiographic markers of left heart function. We used general linear models to examine associations between each exposure and outcome variable.

**Results:** In 1,913 participants with cognitive testing and echocardiography data, we found no associations between any of the echocardiographic exposure variables and worse scores on any of the cognitive tests. Similarly, in 1,035 participants with echocardiography and MRI data, we found no associations between any of the echocardiographic exposure variables and any of the MRI variables.

**Discussion:** We found no associations in this cohort of right heart structure and function with cognition or brain MRI measures. These findings do not support the hypothesis that right heart dysfunction impedes venous return sufficiently to adversely affect brain health among people without clinically apparent heart failure.

---

Cardiovascular disease and dementia rank among the most common causes of disability.^1^ Several cardiac diseases, including coronary artery disease,^2^ heart failure,^3^ and atrial fibrillation,^4^ have been established as risk factors for vascular brain injury and dementia. Subclinical abnormalities of the structure and function of the left heart^5-7^ are associated with vascular brain injury and dementia even in the absence of clinically apparent cardiac disease. The mechanisms underlying these associations are thought to include shared vascular risk factors,^8^ decreased cardiac output,^9^ and cardiac embolism.^10^

Two studies found that elevated right atrial (RA) and jugular venous pressure were associated with regional and total cerebral white matter disease measures,^11,12^ raising the possibility that the right heart function may affect brain health, potentially by impairing venous return from the brain or through other mechanisms. These prior studies involved highly selected patient populations such as patients undergoing right heart catheterization and lacked adjustment for left heart function. To explore these potential connections, we examined the association of right heart structure and function with cognitive function and MRI markers of brain health in the Multi-Ethnic Study of Atherosclerosis (MESA).

## Methods

### Design

MESA is a prospective, longitudinal cohort study focused on subclinical and clinical cardiovascular disease. Baseline examinations took place from 2000 to 2002 and Exam 6 occurred between 2016 and 2018. All participants provided written, informed consent for participation in MESA, and institutional review boards at all participating sites approved this study. Further details of the MESA study have been published previously.^13^

### Participants

MESA enrolled 6,814 men and women 45 to 84 years of age who at baseline in 2000-2002 were free of clinically apparent cerebrovascular or cardiovascular disease. For this analysis, we excluded MESA participants with an adjudicated stroke prior to Exam 6. All other participants with available exposure and outcome data, as defined below, were included in our analyses.

### Measurements

The exposure variables were ascertained from study echocardiograms performed at MESA Exam 6 using standard methods.^14^ These variables included RA pressure, RA end-systolic area, right ventricular (RV) free wall strain, RV fractional area change, RV end-diastolic area, and pulmonary artery systolic pressure. RA pressure was estimated from the diameter and collapsibility of the inferior vena cava during respiration or after a ‘sniff’. RA end-systolic area was traced at the end of ventricular systole by following the RA endocardium from the lateral aspect of the tricuspid annulus to the septal aspect, excluding the area between leaflets and annulus, the inferior vena cava, superior vena cava, and RA appendage. RV free wall strain was assessed using speckle-tracking echocardiography and was computed as the average of the peak longitudinal strain values of the three RV free wall segments. RV areas at end systole and end diastole were assessed using the apical 4-chamber view, tracing from the lateral tricuspid annulus, along the free wall to the apex, and then back to the medial annulus, along the interventricular septum; RV trabeculations, papillary muscles, and the moderator band were included in the cavity area. RV fractional area change was calculated as: (RV end-diastolic area – RV end-systolic area) x 100% / RV end-diastolic area. The pulmonary artery systolic pressure was estimated based on measurement of the velocity of the tricuspid regurgitant jet (V_TR_) and the following formula: 4(V_TR_)^2^ + RA pressure.

The outcome variables included the core MESA cognitive tests from Exam 6: Cognitive Abilities Screening Instrument (CASI; Version 2), Digit Symbol Coding, and Digit Span total (forward and backward).^15^ We excluded 197 participants’ scores on the CASI that were flagged as invalid. To examine antecedent cognitive performance and changes in cognition over time, we assessed the cognitive tests above from Exam 5 (2010-2012) and the change in these tests from Exam 5 to 6. We assessed the following brain MRI variables from scans completed in 2018-19, a median of 18 months after Exam 6: volumes of total gray matter, total white matter, and white matter hyperintensity, all calculated as a percentage of intracranial volume, as well as total white matter fractional anisotropy. The latter is a measure of white matter microstructural integrity, which is decreased in small-vessel cerebrovascular disease. Technical details of the acquisition, processing, and interpretation of these images have been previously published.^16^

Covariates were demographics (age, sex-gender, race-ethnicity, education, and field center), *APOE*-ε4 allele genotype, vascular risk factors measured at Exam 6 (systolic blood pressure, use of antihypertensive medication, diabetes mellitus, body mass index, smoking status, and history of atrial fibrillation, myocardial infarction, and heart failure), and echocardiographic markers of left heart function (left ventricular [LV] ejection fraction, LV mass, LV global longitudinal strain, E/e’ ratio, left atrial [LA] volume, and LA reservoir strain.

### Statistical Analysis

We performed cross-sectional analyses of right heart variables with cognitive test results and brain MRI variables. We used general linear models to examine associations between each exposure variable and each outcome variable. Model 1 included age, sex, race-ethnicity, education, study site, and *APOE*-ε4 allele genotype. Model 2 additionally included systolic blood pressure, use of antihypertensive medication, diabetes mellitus, body mass index, smoking status, and prevalent atrial fibrillation, myocardial infarction, and clinical heart failure. Model 3 additionally included LV ejection fraction, LV mass, LA volume, and E/e’ ratio. We performed subgroup analyses stratified by age (<75 versus ≥75 years), sex, race (White versus non-White), and *APOE*-ε4 allele status (carrier versus not).

## Results

Among 3,303 MESA participants who returned for Exam 6, 103 (3.1%) had a prevalent stroke and were excluded. Of the remaining 3,200 participants, 1,913 (59.8%) had concurrent echocardiographic and cognitive testing data at Exam 6, and 1,035 (32.3%) had both echocardiographic and brain volume data. Compared to those with missing data, participants included in our analyses tended to be younger, had different distributions of race and ethnicity, and had a somewhat lower burden of vascular risk factors (Tables 1 and 2).

**Table 1.** Characteristics of MESA Participants without Prevalent Stroke at Exam 6, Stratified by Availability of Echocardiogram and Cognitive Test Data.

| Characteristic<sup>a</sup> | Echocardiogram and<br>Cognitive Tests<br>Available<br>(N = 1,913) | Echocardiogram and<br>Cognitive Tests<br>Not Available<br>(N = 1,287) |
| --- | --- | --- |
| Age, mean (SD), y | 73.6 (8.4) | 74.8 (9.0) |
| Female | 1,005 (52.5) | 704 (54.7) |
| Race-ethnicity |  |  |
| White | 810 (42.3) | 477 (37.1) |
| Black | 541 (28.3) | 277 (21.5) |
| Hispanic/Latino | 340 (17.8) | 342 (26.6) |
| Chinese | 222 (11.6) | 191 (14.8) |
| Education |  |  |
| Less than 12th grade | 207 (10.8) | 204 (15.9) |
| Completed high school | 771 (40.3) | 513 (39.9) |
| Associate, bachelor, or professional degree | 935 (48.9) | 570 (44.3) |
| Apolipoprotein E genotype |  |  |
| e2/e4 | 54 (3.0) | 30 (2.5) |
| e3/e4 | 394 (21.8) | 232 (19.4) |
| e4/e4 | 38 (2.1) | 27 (2.3) |
| Diabetes status |  |  |
| Normal | 1,018 (54.0) | 585 (49.7) |
| Impaired fasting glucose | 447 (23.7) | 267 (22.7) |
| Untreated diabetes | 78 (4.1) | 43 (3.7) |
| Treated diabetes | 344 (18.2) | 283 (24.0) |
| Cigarette smoking status |  |  |
| Never | 892 (46.7) | 586 (45.8) |
| Former | 910 (47.6) | 625 (48.8) |
| Current | 109 (5.7) | 69 (5.4) |
| Antihypertensive medication | 1,157 (60.8) | 793 (62.1) |
| Atrial fibrillation | 209 (10.9) | 133 (10.3) |
| Myocardial infarction | 46 (2.4) | 46 (3.6) |
| Heart failure | 39 (2.0) | 33 (2.6) |
| Abbreviations: MESA, Multi-Ethnic Study of Atherosclerosis; SD, standard deviation. |  |  |
| <sup>a</sup>Data are presented as number (percent) unless otherwise noted. |  |  |

**Table 2.**
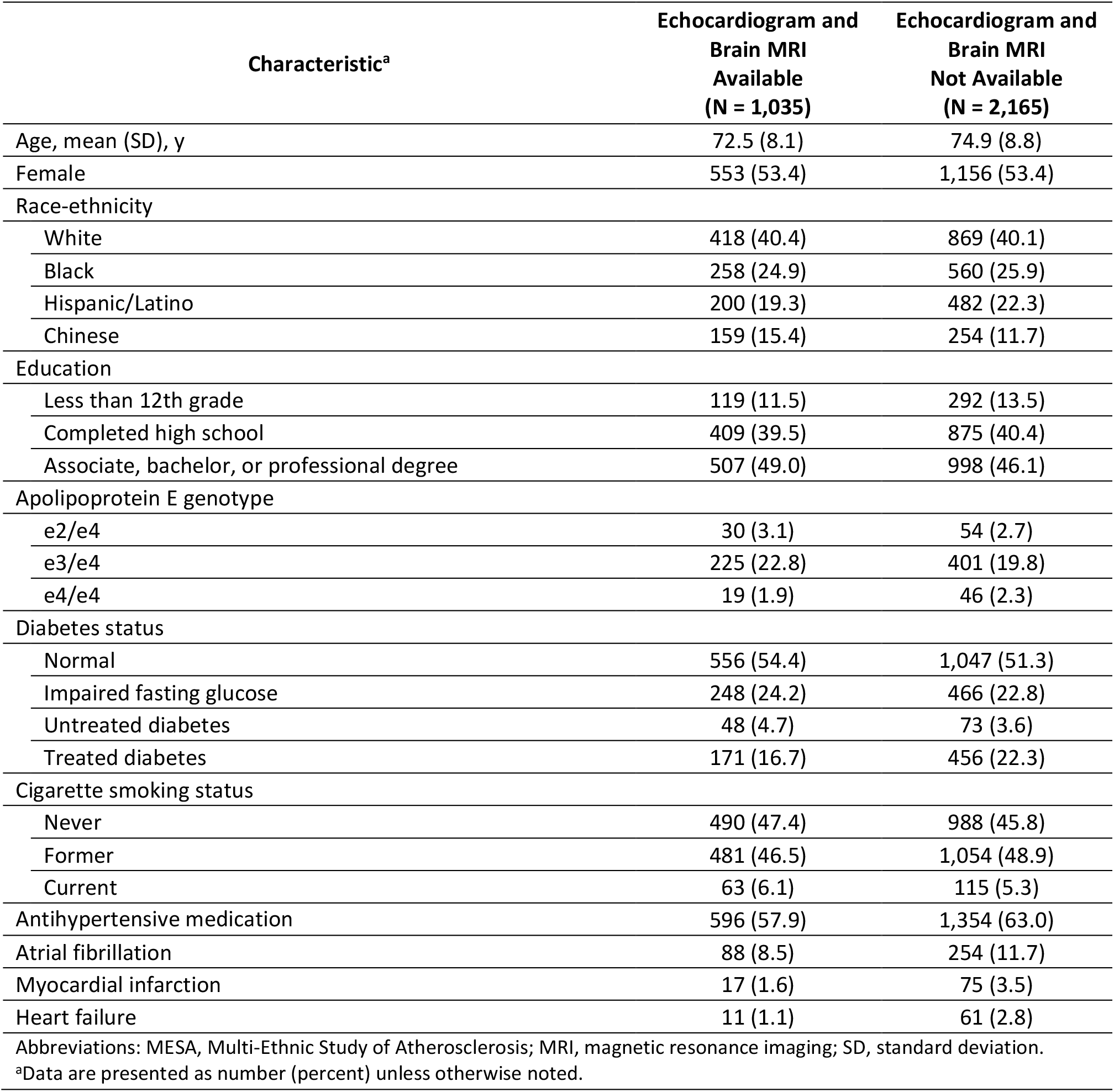
Characteristics of MESA Participants without Prevalent Stroke at Exam 6, Stratified by Availability of Echocardiogram and Brain MRI Data.

| Characteristic<sup>a</sup> | Echocardiogram and<br>Brain MRI<br>Available<br>(N = 1,035) | Echocardiogram and<br>Brain MRI<br>Not Available<br>(N = 2,165) |
| --- | --- | --- |
| Age, mean (SD), y | 72.5 (8.1) | 74.9 (8.8) |
| Female | 553 (53.4) | 1,156 (53.4) |
| Race-ethnicity |  |  |
| White | 418 (40.4) | 869 (40.1) |
| Black | 258 (24.9) | 560 (25.9) |
| Hispanic/Latino | 200 (19.3) | 482 (22.3) |
| Chinese | 159 (15.4) | 254 (11.7) |
| Education |  |  |
| Less than 12th grade | 119 (11.5) | 292 (13.5) |
| Completed high school | 409 (39.5) | 875 (40.4) |
| Associate, bachelor, or professional degree | 507 (49.0) | 998 (46.1) |
| Apolipoprotein E genotype |  |  |
| e2/e4 | 30 (3.1) | 54 (2.7) |
| e3/e4 | 225 (22.8) | 401 (19.8) |
| e4/e4 | 19 (1.9) | 46 (2.3) |
| Diabetes status |  |  |
| Normal | 556 (54.4) | 1,047 (51.3) |
| Impaired fasting glucose | 248 (24.2) | 466 (22.8) |
| Untreated diabetes | 48 (4.7) | 73 (3.6) |
| Treated diabetes | 171 (16.7) | 456 (22.3) |
| Cigarette smoking status |  |  |
| Never | 490 (47.4) | 988 (45.8) |
| Former | 481 (46.5) | 1,054 (48.9) |
| Current | 63 (6.1) | 115 (5.3) |
| Antihypertensive medication | 596 (57.9) | 1,354 (63.0) |
| Atrial fibrillation | 88 (8.5) | 254 (11.7) |
| Myocardial infarction | 17 (1.6) | 75 (3.5) |
| Heart failure | 11 (1.1) | 61 (2.8) |
| Abbreviations: MESA, Multi-Ethnic Study of Atherosclerosis; MRI, magnetic resonance imaging; SD, standard deviation. |  |  |
| <sup>a</sup>Data are presented as number (percent) unless otherwise noted. |  |  |

Among participants included in our analysis of cognition, mean (±SD) scores were 90.1 (±7.5) for the CASI (out of 100), 15.2 (±4.4) for Digit Span total (out of 30), and 50.6 (±18.2) for Digit Symbol Coding (out of 133). Mean (±SD) echocardiographic values were 5.3 mm Hg (±1.4) for RA pressure, 32.9 mm Hg (±7.5) for pulmonary artery systolic pressure, 16.8 cm^2^ (±4.2) for RA end-systolic area, 18.7 cm^2^ (±4.1) for RV end-diastolic area, 40.0% (±4.5) for RV fractional area change, and 24.6 (±5.0) for RV free wall strain. In models adjusted for demographics, we found no significant associations between any of our echocardiographic exposure variables and worse scores on the CASI, Digit Span, or Digit Symbol Coding tests (Table 3). Relationships did not differ notably in progressively adjusted models and were consistent across subgroups. We similarly found no associations between echocardiographic variables and cognitive performance at Exam 5 (Table 4) or greater worsening from Exam 5 to Exam 6 (Table 5).

**Table 3.** Cross-Sectional Associations of Right Heart Structure and Function with Cognitive Test Scores in the Multi-Ethnic Study of Atherosclerosis.

| Cognitive Test | Model 1 | Model 2 | Model 3 |
| --- | --- | --- | --- |
| **CASI** |  |  |  |
| RA pressure | 0.24 (-0.06, 0.54) | 0.19 (-0.11, 0.50) | 0.36 (0.04, 0.69) |
| PA systolic pressure | 0.18 (-0.21, 0.56) | 0.33 (-0.06, 0.72) | 0.42 (0.01, 0.84) |
| RA area | -0.25 (-0.58, 0.07) | -0.25 (-0.59, 0.09) | -0.25 (-0.64, 0.14) |
| RV end diastolic area | -0.28 (-0.64, 0.08) | -0.26 (-0.64, 0.12) | -0.19 (-0.59, 0.22) |
| RV fractional area change | 0.20 (-0.10, 0.49) | 0.20 (-0.09, 0.49) | 0.25 (-0.06, 0.56) |
| RV free wall strain | -0.19 (-0.50, 0.13) | -0.07 (-0.39, 0.25) | -0.04 (-0.38, 0.29) |
| **Digit Span** |  |  |  |
| RA pressure | 0.12 (-0.05, 0.30) | 0.09 (-0.09, 0.27) | 0.17 (-0.02, 0.36) |
| PA systolic pressure | 0.13 (-0.08, 0.34) | 0.14 (-0.09, 0.36) | 0.18 (-0.06, 0.41) |
| RA area | -0.20 (-0.39, 0.00) | -0.16 (-0.37, 0.04) | -0.15 (-0.39, 0.08) |
| RV end diastolic area | -0.12 (-0.33, 0.10) | -0.05 (-0.28, 0.18) | 0.02 (-0.23, 0.27) |
| RV fractional area change | 0.05 (-0.12, 0.23) | 0.03 (-0.15, 0.21) | 0.03 (-0.16, 0.21) |
| RV free wall strain | -0.10 (-0.29, 0.08) | -0.16 (-0.35, 0.03) | -0.16 (-0.36, 0.04) |
| **Digit Symbol Coding** |  |  |  |
| RA pressure | 0.16 (-0.47, 0.80) | 0.15 (-0.50, 0.80) | 0.25 (-0.42, 0.93) |
| PA systolic pressure | -0.26 (-1.03, 0.51) | 0.13 (-0.67, 0.93) | 0.26 (-0.58, 1.10) |
| RA area | 0.08 (-0.62, 0.78) | 0.44 (-0.31, 1.19) | 0.65 (-0.20, 1.50) |
| RV end diastolic area | -0.35 (-1.13, 0.43) | 0.20 (-0.64, 1.04) | 0.46 (-0.43, 1.34) |
| RV fractional area change | 0.36 (-0.27, 0.99) | 0.22 (-0.41, 0.86) | 0.35 (-0.31, 1.01) |
| RV free wall strain | 0.43 (-0.24, 1.10) | 0.32 (-0.38, 1.01) | 0.27 (-0.45, 0.98) |

Abbreviations: CASI, Cognitive Assessment Screening Instrument; PA, pulmonary artery; RA, right atrial; RV, right ventricular.
Data are presented as adjusted mean differences (95% confidence intervals) in cognitive test scores per standard-deviation increase in echocardiographic exposure variables. Echocardiographic and cognitive testing variables were both from Exam 6 (2016-2018). Model 1 included age, sex, race-ethnicity, education, study site, and *APOE*- ε4 allele status. Model 2 additionally included systolic blood pressure, use of antihypertensive medication, diabetes mellitus, body mass index, smoking status, and prevalent atrial fibrillation, myocardial infarction, and clinical heart failure. Model 3 additionally included left ventricular ejection fraction, left ventricular mass, left atrial volume, and E/e' ratio.

**Table 4.** Associations of Right Heart Structure and Function with Antecedent Cognitive Test Scores in the Multi-Ethnic Study of Atherosclerosis.

| **Cognitive Test** | **Model 1** | **Model 2** | **Model 3** |
| --- | --- | --- | --- |
| **CASI** |  |  |  |
| RA pressure | 0.24 (0.00, 0.47) | 0.17 (-0.08, 0.41) | 0.16 (-0.09, 0.42) |
| PA systolic pressure | 0.18 (-0.11, 0.46) | 0.19 (-0.11, 0.49) | 0.26 (-0.06, 0.57) |
| RA area | 0.03 (-0.23, 0.29) | 0.00 (-0.28, 0.27) | -0.05 (-0.37, 0.27) |
| RV end diastolic area | 0.25 (-0.04, 0.54) | 0.34 (0.03, 0.66) | 0.33 (0.00, 0.67) |
| RV fractional area change | 0.06 (-0.18, 0.31) | 0.10 (-0.15, 0.35) | 0.10 (-0.15, 0.36) |
| RV free wall strain | -0.03 (-0.28, 0.21) | -0.02 (-0.29, 0.24) | -0.02 (-0.29, 0.25) |
| **Digit Span** |  |  |  |
| RA pressure | -0.03 (-0.18, 0.12) | -0.05 (-0.21, 0.11) | -0.04 (-0.21, 0.13) |
| PA systolic pressure | 0.10 (-0.09, 0.28) | 0.12 (-0.07, 0.31) | 0.15 (-0.05, 0.36) |
| RA area | -0.13 (-0.30, 0.04) | -0.08 (-0.26, 0.10) | -0.08 (-0.29, 0.13) |
| RV end diastolic area | -0.02 (-0.21, 0.17) | 0.08 (-0.12, 0.29) | 0.13 (-0.09, 0.35) |
| RV fractional area change | 0.03 (-0.13, 0.19) | -0.01 (-0.17, 0.15) | -0.01 (-0.18, 0.15) |
| RV free wall strain | -0.10 (-0.26, 0.06) | -0.21 (-0.38, -0.04) | -0.21 (-0.38, -0.03) |
| **Digit Symbol Coding** |  |  |  |
| RA pressure | 0.31 (-0.24, 0.85) | 0.43 (-0.12, 0.98) | 0.48 (-0.09, 1.06) |
| PA systolic pressure | -0.20 (-0.82, 0.42) | 0.04 (-0.61, 0.68) | 0.05 (-0.63, 0.73) |
| RA area | -0.30 (-0.90, 0.29) | -0.09 (-0.72, 0.53) | -0.40 (-1.13, 0.33) |
| RV end diastolic area | -0.24 (-0.91, 0.42) | 0.19 (-0.52, 0.90) | 0.14 (-0.62, 0.90) |
| RV fractional area change | 0.18 (-0.37, 0.72) | 0.02 (-0.53, 0.57) | 0.12 (-0.45, 0.69) |
| RV free wall strain | 0.20 (-0.35, 0.75) | -0.09 (-0.67, 0.49) | -0.16 (-0.75, 0.44) |

Abbreviations: CASI, Cognitive Assessment Screening Instrument; PA, pulmonary artery; RA, right atrial; RV, right ventricular.
Data are presented as adjusted mean differences (95% confidence intervals) in cognitive test scores per standard-deviation increase in echocardiographic exposure variables. Cognitive testing variables were from Exam 5 (2010-2012) and echocardiographic data were from Exam 6 (2016-2018). Model 1 included age, sex, race-ethnicity, education, study site, and *APOE*- ε4 allele status. Model 2 additionally included systolic blood pressure, use of antihypertensive medication, diabetes mellitus, body mass index, smoking status, and prevalent atrial fibrillation, myocardial infarction, and clinical heart failure. Model 3 additionally included left ventricular ejection fraction, left ventricular mass, left atrial volume, and E/e' ratio.

**Table 5.** Associations of Right Heart Structure and Function with Change in Cognitive Test Scores in the Multi-Ethnic Study of Atherosclerosis.

| **Cognitive Test** | **Model 1** | **Model 2** | **Model 3** |
| --- | --- | --- | --- |
| **CASI** |  |  |  |
| RA pressure | -0.01 (-0.32, 0.31) | 0.05 (-0.28, 0.37) | 0.20 (-0.15, 0.54) |
| PA systolic pressure | -0.05 (-0.44, 0.34) | 0.02 (-0.38, 0.43) | 0.03 (-0.39, 0.46) |
| RA area | -0.24 (-0.59, 0.10) | -0.26 (-0.62, 0.10) | -0.34 (-0.76, 0.07) |
| RV end diastolic area | -0.34 (-0.72, 0.04) | -0.42 (-0.82, -0.02) | -0.41 (-0.84, 0.02) |
| RV fractional area change | 0.10 (-0.21, 0.41) | 0.08 (-0.23, 0.39) | 0.09 (-0.24, 0.42) |
| RV free wall strain | -0.03 (-0.36, 0.30) | 0.10 (-0.24, 0.44) | 0.07 (-0.29, 0.42) |
| **Digit Span** |  |  |  |
| RA pressure | 0.14 (-0.01, 0.29) | 0.13 (-0.03, 0.29) | 0.17 (0.01, 0.34) |
| PA systolic pressure | 0.03 (-0.15, 0.21) | 0.04 (-0.16, 0.23) | 0.02 (-0.19, 0.23) |
| RA area | -0.03 (-0.20, 0.14) | -0.08 (-0.26, 0.10) | -0.08 (-0.29, 0.14) |
| RV end diastolic area | 0.02 (-0.16, 0.21) | -0.03 (-0.24, 0.17) | -0.03 (-0.24, 0.19) |
| RV fractional area change | -0.07 (-0.22, 0.08) | -0.04 (-0.19, 0.11) | -0.06 (-0.22, 0.11) |
| RV free wall strain | -0.03 (-0.20, 0.13) | 0.01 (-0.16, 0.18) | 0.02 (-0.16, 0.20) |
| **Digit Symbol Coding** |  |  |  |
| RA pressure | 0.11 (-0.47, 0.69) | -0.03 (-0.62, 0.57) | 0.05 (-0.57, 0.67) |
| PA systolic pressure | -0.16 (-0.85, 0.54) | -0.12 (-0.85, 0.62) | -0.01 (-0.78, 0.77) |
| RA area | 0.46 (-0.19, 1.10) | 0.48 (-0.20, 1.17) | 0.96 (0.17, 1.75) |
| RV end diastolic area | 0.18 (-0.53, 0.90) | 0.25 (-0.52, 1.02) | 0.54 (-0.29, 1.37) |
| RV fractional area change | 0.20 (-0.36, 0.76) | 0.28 (-0.30, 0.85) | 0.29 (-0.32, 0.89) |
| RV free wall strain | 0.41 (-0.19, 1.02) | 0.56 (-0.07, 1.19) | 0.60 (-0.06, 1.26) |

Abbreviations: CASI, Cognitive Assessment Screening Instrument; PA, pulmonary artery; RA, right atrial; RV, right ventricular.
Data are presented as adjusted mean differences (95% confidence intervals) in cognitive test score changes from Exam 5 (2010-2012) to Exam 6 (2016-2018) per standard-deviation increase in echocardiographic exposure variables at Exam 6. Model 1 included age, sex, race-ethnicity, education, study site, and *APOE*- ε4 allele status. Model 2 additionally included systolic blood pressure, use of antihypertensive medication, diabetes mellitus, body mass index, smoking status, and prevalent atrial fibrillation, myocardial infarction, and clinical heart failure. Model 3 additionally included left ventricular ejection fraction, left ventricular mass, left atrial volume, and E/e' ratio.

Among participants included in our analysis of brain imaging, mean (±SD) volumes were 0.5% (±0.7) of intracranial volume for white matter hyperintensity, 36.5% (±1.7) for total white matter, and 44.0% (±3.0) for total gray matter. Mean (±SD) total white matter fractional anisotropy was 0.39 (±0.03). Mean echocardiographic values were 5.2 mm Hg (±1.1) for RA pressure, 32.6 mm Hg (±6.6) for pulmonary artery systolic pressure, 16.5 cm^2^ (±4.0) for RA end-systolic area, 18.4 cm^2^ (±4.0) for RV end-diastolic area, 40.1% (±4.4) for RV fractional area change, and 24.9 (±5.0) for RV free wall strain. We found no significant associations between any of our echocardiographic exposure variables and white matter hyperintensity, white matter, or gray matter volumes or total white matter fractional anisotropy (Table 6). Again, associations did not differ notably in progressively adjusted models.

**Table 6.** Cross-Sectional Associations of Right Heart Structure and Function with Brain MRI Measures in the Multi-Ethnic Study of Atherosclerosis.

| Brain MRI Variable | Model 1 | Model 2 | Model 3 |
| --- | --- | --- | --- |
| White matter hyperintensity volume |  |  |  |
| RA pressure | -0.01 (-0.05, 0.03) | -0.01 (-0.05, 0.04) | -0.01 (-0.06, 0.04) |
| PA systolic pressure | 0.01 (-0.04, 0.06) | 0.00 (-0.05, 0.05) | -0.01 (-0.07, 0.04) |
| RA area | 0.00 (-0.05, 0.04) | 0.00 (-0.05, 0.05) | 0.01 (-0.05, 0.07) |
| RV end diastolic area | 0.00 (-0.05, 0.06) | 0.00 (-0.06, 0.06) | 0.00 (-0.06, 0.07) |
| RV fractional area change | -0.05 (-0.09, 0.00) | -0.05 (-0.10, -0.01) | -0.06 (-0.11, -0.01) |
| RV free wall strain | -0.04 (-0.09, 0.00) | -0.05 (-0.10, 0.00) | -0.05 (-0.10, 0.01) |
| White matter volume |  |  |  |
| RA pressure | -0.03 (-0.13, 0.07) | -0.03 (-0.14, 0.07) | -0.05 (-0.15, 0.06) |
| PA systolic pressure | 0.03 (-0.09, 0.15) | 0.03 (-0.09, 0.15) | 0.01 (-0.12, 0.14) |
| RA area | 0.00 (-0.11, 0.11) | 0.03 (-0.09, 0.15) | 0.04 (-0.10, 0.17) |
| RV end diastolic area | -0.05 (-0.17, 0.08) | -0.02 (-0.15, 0.12) | 0.01 (-0.14, 0.16) |
| RV fractional area change | 0.03 (-0.07, 0.14) | 0.02 (-0.09, 0.12) | 0.02 (-0.09, 0.13) |
| RV free wall strain | 0.01 (-0.10, 0.12) | -0.02 (-0.13, 0.09) | 0.00 (-0.12, 0.12) |
| Gray matter volume |  |  |  |
| RA pressure | -0.06 (-0.20, 0.08) | -0.03 (-0.17, 0.12) | -0.01 (-0.15, 0.14) |
| PA systolic pressure | -0.02 (-0.18, 0.15) | 0.01 (-0.16, 0.18) | 0.01 (-0.16, 0.19) |
| RA area | -0.02 (-0.17, 0.14) | 0.03 (-0.13, 0.19) | 0.00 (-0.18, 0.18) |
| RV end diastolic area | 0.00 (-0.17, 0.18) | 0.04 (-0.15, 0.23) | 0.02 (-0.19, 0.22) |
| RV fractional area change | 0.12 (-0.02, 0.27) | 0.11 (-0.04, 0.25) | 0.07 (-0.08, 0.22) |
| RV free wall strain | 0.13 (-0.02, 0.28) | 0.11 (-0.05, 0.26) | 0.10 (-0.05, 0.26) |
| Fractional anisotropy |  |  |  |
| RA pressure | -0.06 (-0.20, 0.09) | -0.07 (-0.22, 0.07) | -0.04 (-0.19, 0.11) |
| PA systolic pressure | -0.06 (-0.22, 0.11) | -0.01 (-0.17, 0.16) | 0.04 (-0.14, 0.21) |
| RA area | -0.08 (-0.24, 0.07) | -0.01 (-0.17, 0.15) | 0.01 (-0.17, 0.20) |
| RV end diastolic area | -0.05 (-0.23, 0.13) | 0.10 (-0.09, 0.29) | 0.12 (-0.08, 0.32) |
| RV fractional area change | 0.11 (-0.03, 0.26) | 0.09 (-0.05, 0.24) | 0.12 (-0.03, 0.27) |
| RV free wall strain | 0.15 (0.00, 0.31) | 0.10 (-0.06, 0.26) | 0.10 (-0.06, 0.27) |

Abbreviations: PA, pulmonary artery; RA, right atrial; RV, right ventricular.
Data are presented as adjusted mean differences (95% confidence intervals) in MRI measures per standard-deviation increase in echocardiographic exposure variables. Echocardiographic variables were from Exam 6 (2016-2018) and brain volume variables were from 2018-2019. All brain volumes were modeled as percent of intracranial volume. Fractional anisotropy was multiplied by 100 to enable more easily interpreted adjusted mean differences. Model 1 included age, sex, race-ethnicity, education, study site, and *APOE*- ε4 allele status. Model 2 additionally included systolic blood pressure, use of antihypertensive medication, diabetes mellitus, body mass index, smoking status, and prevalent atrial fibrillation, myocardial infarction, and clinical heart failure. Model 3 additionally included left ventricular ejection fraction, left ventricular mass, left atrial volume, and E/e' ratio.

## Discussion

Using data from a longitudinal cohort study of a diverse community-dwelling sample of older adults, we found that echocardiographic measurements of right heart structure and function were not associated with cognitive function or MRI measures of brain health. The null findings were consistent across numerous exposure-outcome pairs, were consistent across subgroups, and did not change appreciably with or without adjustment for demographics, vascular risk factors, and echocardiographic measurements of left heart structure and function.

Although numerous prior studies have examined the association of left heart structure and function with cognition and brain MRI measures,^5-7^ published data on relationships between the right heart and brain health are sparse. Studies have found that decreased clearance of amyloid and other toxic substances from the brain may play a role in Alzheimer’s Disease and related dementias.^17^ Since the venous system is a vital clearance pathway for cerebrospinal fluid and disruption in venous drainage alters the cerebral white matter, impaired venous return to the heart may have a negative impact brain health.^18^ Two prior studies found that elevated venous pressure was associated with cerebral white matter disease, but these involved relatively small and highly selected cohorts.^11,12^ In this context, our study adds novel data on the relationship of right heart structure and function with cognition and brain volumes in a relatively large, community-based cohort. Our findings do not support the hypothesis that alterations in right heart structure and function affect venous return from the brain to a sufficient degree to affect brain health, at least among relatively healthy individuals without clinically manifest heart failure.

Our study has several limitations. First, it involves a relatively healthy cohort of individuals free of clinically apparent vascular disease at baseline. Participants who returned for Exam 6 and had available echocardiographic and cognitive testing data were even healthier at baseline than the overall MESA cohort. Few participants had notably abnormal right heart structure or function, so these results leave open the possibility that more extreme right heart dysfunction may adversely affect brain health. Our cohort was generally younger, healthier, and had fewer right heart abnormalities than patients in the prior studies, which reported associations between elevated RA pressure and white matter disease. Nevertheless, prior studies have shown associations between mostly subclinical left heart dysfunction and brain health in MESA,^7^ so our findings may still provide valuable insight into brain health across the spectrum of right heart function and subclinical dysfunction in this same cohort. Second, our cross-sectional design may have missed long-term effects of abnormal right heart structure and function on cognition. Third, we only had noninvasive, indirect measurements of RA pressure, and possibly direct measurements of RA pressure may have revealed associations with cognition. Fourth, the cognitive data currently available in MESA are limited to a few tests, and more comprehensive testing may have revealed associations between specific cognitive domains, or impairment, and our echocardiographic variables.

In summary, we found no associations between echocardiographic measurements of right heart structure and function and measurements of cognition or MRI measures of brain health. These findings do not support the hypothesis that variations in right heart function outside the context of clinically apparent heart failure impedes venous return sufficiently to adversely affect brain health.

## Data Availability

All data produced in the present study are available upon reasonable request to the authors.

## Acknowledgements

None.

